# Tandem standing duration is a rapid, sensitive and specific test of Parkinson’s Disease subtype

**DOI:** 10.1101/2024.07.19.24310694

**Authors:** Sarah Hosli, Matteo Ciocca, Zaeem Hadi, Sophie Molloy, Yen Tai, Barry M Seemungal

## Abstract

**Background:** Parkinson’s Disease (PD) patients with postural instability and gait disorder (PIGD) subtype are at increased risk for falls compared to the tremor-dominant subtype. We aimed to establish an easy clinical balance tool to rapidly and reliably identify PIGD patients, potentially important for directing healthcare resources or research phenotyping.

**Methods:** 45 consecutive patients with PD completed clinical testing including Romberg, tandem stance, single leg stance, 360° turning and 10-meter walking. MDS-UPDRS part II and III, collected as part of regular follow-up, was used to classify disease subtype. Multinominal logistic regression models were fitted to find optimal subtype predictors and compared using receiver operating characteristic (ROC) curves.

**Results:** Unassisted tandem stand duration and time to turn 360° were significantly different between PIGD and tremor dominant subtypes. Both tandem standing and 360° turning showed very high predictive accuracy to predict PD subtype with an area under the ROC curve (AUC) of 86.6% and 88% respectively, which increased to 91.4% by combining both measures. Optimal cut-off values for identifying PD subtypes were tandem standing less than 20s and 360° turning longer than 6.5s.

**Conclusion:** Tandem stand duration and 360° turning are easy to apply clinical tests that rapidly identify PD patients with PIGD subtype with high sensitivity and specificity. These findings may be useful in the clinic to identify PD patients’ current falls risk or screening for research studies.

**Plain Language Summary:** Balance problems and falls are common in late-stage Parkinson’s Disease, affecting nearly 70% of patients 10 years post-diagnosis. In contrast, Parkinson’s patients who complain mainly of shaking (tremor) are less liable to fall. We set out to find an easy and reliable bedside test to distinguish patients at risk of falls with early Parkinson’s. This is important so that resources can be targeted to patients in need of support such as physiotherapy and fall prevention. 45 patients with Parkinson’s disease participated in this study and completed a battery of balance tests completed within the time of their regular follow-up appointment. We found that tandem standing duration – a test where patient stand still in the heel-to-toe position – and time taken to complete a full circle, were highly reliable in detecting patients with balance and gait problems. Specifically, patients with balance and gait problems were unable to tandem stand for more than 20 seconds and took more than 6.5 seconds to turn a full circle. Together, these two tests that take a minute to complete in the clinic, and may help improve the care for patients with Parkinson’s as a quick screening tool to identify Parkinson’s disease at risk of falls.

## Introduction

Neurological disorders are the leading source of disability worldwide, and in 2016, 6 million individuals were affected by Parkinson’s disease (PD). [1] This number is believed to increase due to a combination of an aging population and external environmental and behavioural factors such as changing smoking patterns or increased exposure to pesticides. [2, 3] Falls are a critical milestone in PD with nearly two-thirds of patients suffering a fall within the first 7 years after their diagnosis, and is a major driver for patient and care-giver socio-economic burden. [4] Within the high-risk group of PD patients, the postural instability and gait difficulties (PIGD) phenotype has been identified as an independent risk factor of falls as well as having a worse prognosis in terms of cognitive and behavioural impairment.[5, 6] An easy clinical screening tool to identify patients at risk of falls, including those that could be undertaken by non-experts, would be important for both the clinic – to identify high risk patients needing additional support and resources – and for research – to rapidly screen for PD subtypes and optimise patient recruitment. Additionally, specific clinical deficits could help stratify patients and identify distinct pathophysiological mechanisms mediating different disease phenotypes. Moreover, both finding treatments for balance problems, and early differentiation of PD subtypes, have been identified as the top three research priorities by Parkinson’s patients and their caregivers in 2014. [7]

Clinical balance testing has been studied to examine different subgroups of healthy participants as well as patients with PD. Tandem stance (heel-to-toe alignment of both feet) proved to be a good tool to assess healthy elderly people for the need of a walking aid. [8] Tandem stance as well as the single leg stance (SLS) are tests with strong test-retest reliability in distinguishing PD patients with and without a history of falls. [9] Tandem stand duration can be used to monitor success of for balance training in PD patients. [10] Other commonly used balance tests include Romberg, 360° turning and evaluation of 10-meter walking speed. While sway analysis of the Romberg Test reveals differences in PD patients compared to healthy controls, clinical testing of the Romberg duration shows a ceiling effect with most participant being able to stand for 60 seconds or longer. [11, 12] 360° turning has been examined as clinical test to predict PD phenotypes, but only showed moderate accuracy when looking at both step count as well as time needed to turn. [13] Lastly, walking speed has been analysed in PIGD and TD PD phenotypes, showing slower walking speeds for PIGD patients as well as a dependence of walking speed on ON or OFF medication state of the participants for both phenotypes. [14]

While prior research has examined different clinical balance tests in the PD population, most of these results showed poor accuracy to determine PD subtypes. The aim of our study was to develop an efficient bedside test to identify patients at risks for balance dysfunction and falls. There has to our knowledge been no comparison of tandem stance alone or in combination with other clinical balance test to determine its predictive ability to distinguish PD phenotypes.

## Methods

### Participants

Participants were recruited from the movement disorders clinic at Imperial College Healthcare NHS Trust, where they were seen as part of a regular follow up. During clinic visits MDS-UPDRS part 2 and 3 were completed. Over 5 months, 200 participants were screened and 45 participants included in the study to undergo further balance tests during their visit. Inclusion criteria were: diagnosis of Parkinson’s disease based on MDS criteria and the ability to stand independently.[15] Exclusion criteria were atypical Parkinson’s disease, patients with non-oral therapies such as deep brain stimulation or apomorphine treatment, patients with a diagnosis of cognitive impairment and patients who relied on walking aids or wheelchair-bound patients who would not be able to complete the testing.

### Protocol

After completing MDS-UPDRS part 2 and 3 during the regular clinic appointment, patients underwent a series of clinical balance tests. Most patients presented in ON-status for the clinical visit and balance examination. However, patients presenting in OFF-status were also included. Clinical ON / OFF status was recorded (as defined in MDS-UPDRS III 3b) for each participant. Romberg test (quiet stance with eyes closed) was performed and recorded over maximum 20 seconds. The time was noted if the patient opened their eyes or took a step before 20s. Tandem stance was recorded three times over maximum period of 30 seconds. Patients were free to choose which foot was placed in front and instructed to align their feet heel to toe and fixate a point in front of them. The time was recorded as soon as the patient took a corrective step to the side. In order to reduce possible confounding, patients were encouraged to get into tandem stance position without any external support.[16] Patients who were not able to get into tandem stance by themselves were recorded as 0 seconds for unassisted tandem stance. For those requiring assistance to assume the tandem stance position, we recorded an assisted tandem stance duration up to 30 seconds for three separate trials. Single-leg stance was performed on the preferred leg and recorded up to a maximum duration of 20 seconds. The time was recorded if the second leg touched the floor before 20s. The duration of 360° turning on the spot was recorded for turns to the right and to the left separately. The time taken for a 10-meter walk was recorded. Patients could use a walking aid for the 10-meter walk if they used one in daily life.

### Statistical Analysis and data availability

Patient data, MDS-UPDRS and balance scores were recorded in patients’ health records. Data analysis was performed using R studio and the nnet and pROC packages. [17, 18] PD subtypes were calculated using MDS-UPDRS scores as described by Stebbins et al, calculating the ratio of TD and PIGD dominant subscores with scores ≥1.15 for TD classification and ≤0.90 for PIGD. Patients with scores between 0.90 and 1.15 were defined as an “indeterminate” group. [19] Multinominal logistic regression models were fitted to predict PD subtype (PIGD, TD or indeterminate), predictors were selected based on model fit using Likelihood Ratio testing. Receiver-Operating Characteristic (ROC) curves were used to evaluate and compare the classification accuracy of the selected predictors and optimal cut-off values identified using Youden’s index.

### Ethics

The study was approved by the UK Health Research Authority (IRAS ID255217; study sponsor Imperial College London), enabling access and analysis of pseudonymized data from routine clinical visits.

## Results

### Clinical and demographic features

45 patients were included in the study (mean age 70, SD 8.7 years). 64% of recruited patients were male, which approximately reflects the gender distribution in the PD population.[20]

After calculating TD and PIGD subscores, 19 patients were classified as TD phenotype, 22 patients as PIGD phenotype and 4 patients as indeterminate. Groups were balanced for gender, disease duration and ON/ OFF medication status, although there is an observation of more patients in OFF status in the PIGD group (22.7%) vs. in the TD group (5.26%). Group differences were found in age and disease severity with PIGD patients being significantly older than TD PD and Indeterminate patients and having higher UPDRS scores. Differences were also seen in Hoehn & Yahr staging, which was to be expected since Hoehn & Yahr stage ≥3 is defined by observation of postural instability.

Results of clinical balance tests were visualized using boxplots to identify potentially useful test to distinguish PD phenotypes *(Figure 1)*. Except for Romberg test, all clinical tests showed different distributions among subgroups. Romberg was tested up to 20 seconds which revealed a clear ceiling effect with all participants but one reaching maximum duration. Tandem stand duration, 360° turning and 10-meter walking were selected for the logistic regression model to predict the PD subtype.

**Figure 1:**
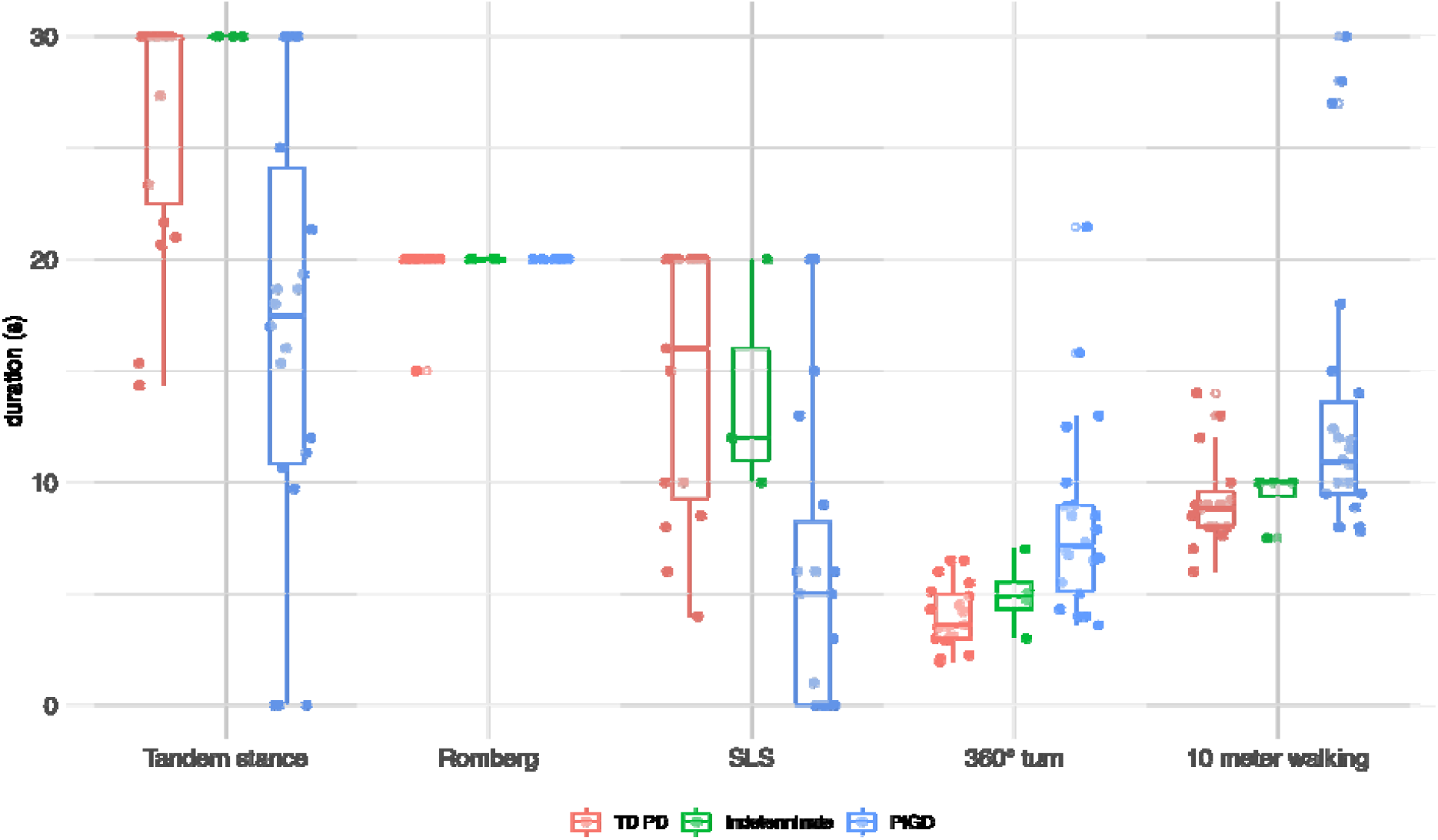
Visualisation of clinical balance test results with boxplots and raw data distribution. SLS = single leg stance

### Multinominal logistic regression

Multinominal logistic regression models were fitted for each clinical balance test. Predictors Age, Sex, MDS-UPDRS, disease duration as well as ON/OFF medication status did not improve model fit. However, age was kept in the final model based on the significant baseline differences that were found between the tested groups. Final models included age and clinical balance test as predictors to determine PD subtype (PIGD, TD or indeterminate), see *Table 2*. Unassisted tandem stand duration adjusted for age showed an odds ratio of 0.87 (*p = 0.016*), with longer tandem stand duration predicting smaller odds of the patient belonging to the PIGD subtype. Time to turn 360° adjusted for age showed an odds ratio of 2.02 (p < 0.01), implying longer time to turn as a predictor for PIGD subtype. Time for 10-meter walking was significant in the unadjusted model (crude OR 1.40 (p= 0.04), but non-significant when adjusting for age, suggesting that walking speed differences between the groups were attributed to the age difference rather than the PD subtype. Because of the expected impact of ON / OFF medication status on some of the balance assessments (such as 360° turning), multinominal logistic regression was repeated adjusted for age and medication status, which did not affect the initial outcome. *(Table 3, supplementary material)*

**Table 1:** Summary descriptives by PD phenotype.

|  | **TD PD** | **Indeterminate** | **PIGD** | **p-value** |
| --- | --- | --- | --- | --- |
|  | *N=19* | *N=4* | *N=22* |  |
| Gender: |  |  |  | 0.424 |
| Female | 9 (47.4%) | 1 (25.0%) | 6 (27.3%) |  |
| Male | 10 (52.6%) | 3 (75.0%) | 16 (72.7%) |  |
| Age (years) | 65.1 (9.33) | 69.9 (9.20) | 73.8 (5.99) | 0.004 |
| Disease duration (years) | 6.83 (4.74) | 6.38 (7.13) | 7.77 (4.60) | 0.772 |
| UPDRS | 19.4 (11.9) | 21.2 (7.41) | 29.4 (11.5) | 0.025 |
| Medication status: |  |  |  | 0.246 |
| ON | 18 (94.7%) | 4 (100%) | 17 (77.3%) |  |
| OFF | 1 (5.26%) | 0 (0.00%) | 5 (22.7%) |  |
| Hoehn & Yahr: |  |  |  | 0.004 |
| 1 | 13 (68.4%) | 1 (25.0%) | 3 (13.6%) |  |
| 2 | 4 (21.1%) | 3 (75.0%) | 14 (63.6%) |  |
| 3 | 2 (10.5%) | 0 (0.00%) | 3 (13.6%) |  |
| 4 | 0 (0.00%) | 0 (0.00%) | 2 (9.09%) |  |

**Table 2:** Multiple logistic regression results for clinical balance tests.

|  | Unadjusted |  | Adjusted<sup>1</sup> |  |
| --- | --- | --- | --- | --- |
|  | OR (95% CI) | p-value | OR (95% CI) | p-value |
| Tandem stand<br>(seconds) | 0.86 (0.75, 0.96) | 0.003** | 0.87 (0.75 to 0.98) | 0.016* |
| 360° turning<br>(seconds) | 2.27 (1.75, 2.80) | 0.002** | 2.02 (1.49, 2.56) | 0.002** |
| 10-meter walking<br>(seconds) | 1.40 (1.08, 1.73) | 0.04* | 1.25 (0.96, 1.55) | 0.13 |

Abbreviations: OR = odds ratio; CI = confidence interval
<sup>1</sup>adjusted for age
\*p < 0.05
\*\*p < 0.01

Supplementary material:

**Table 3:**
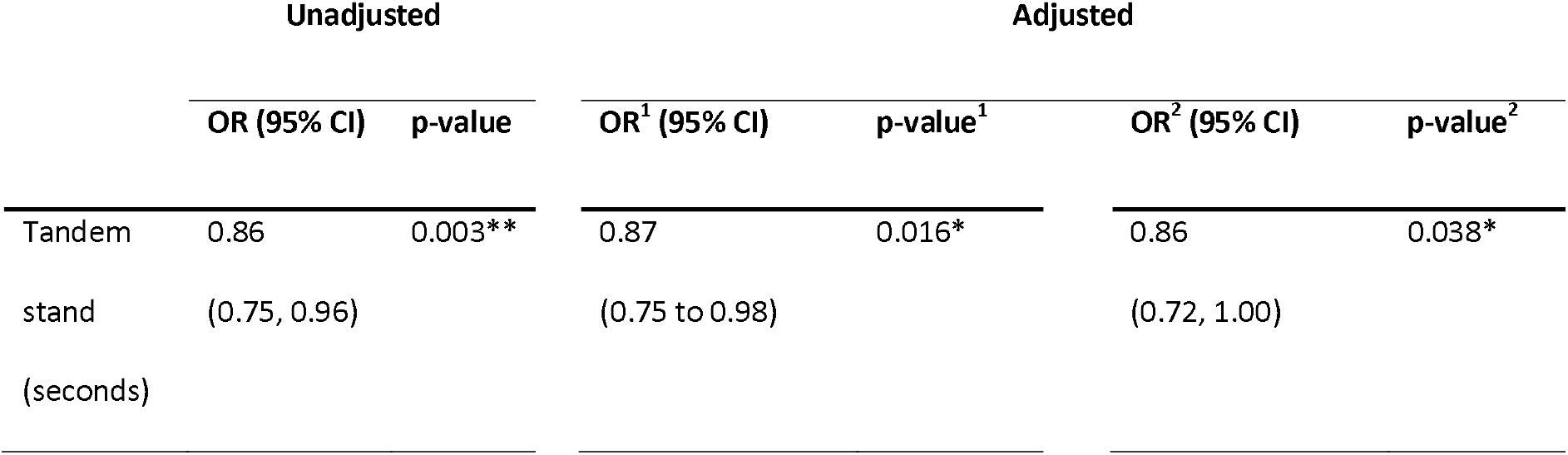
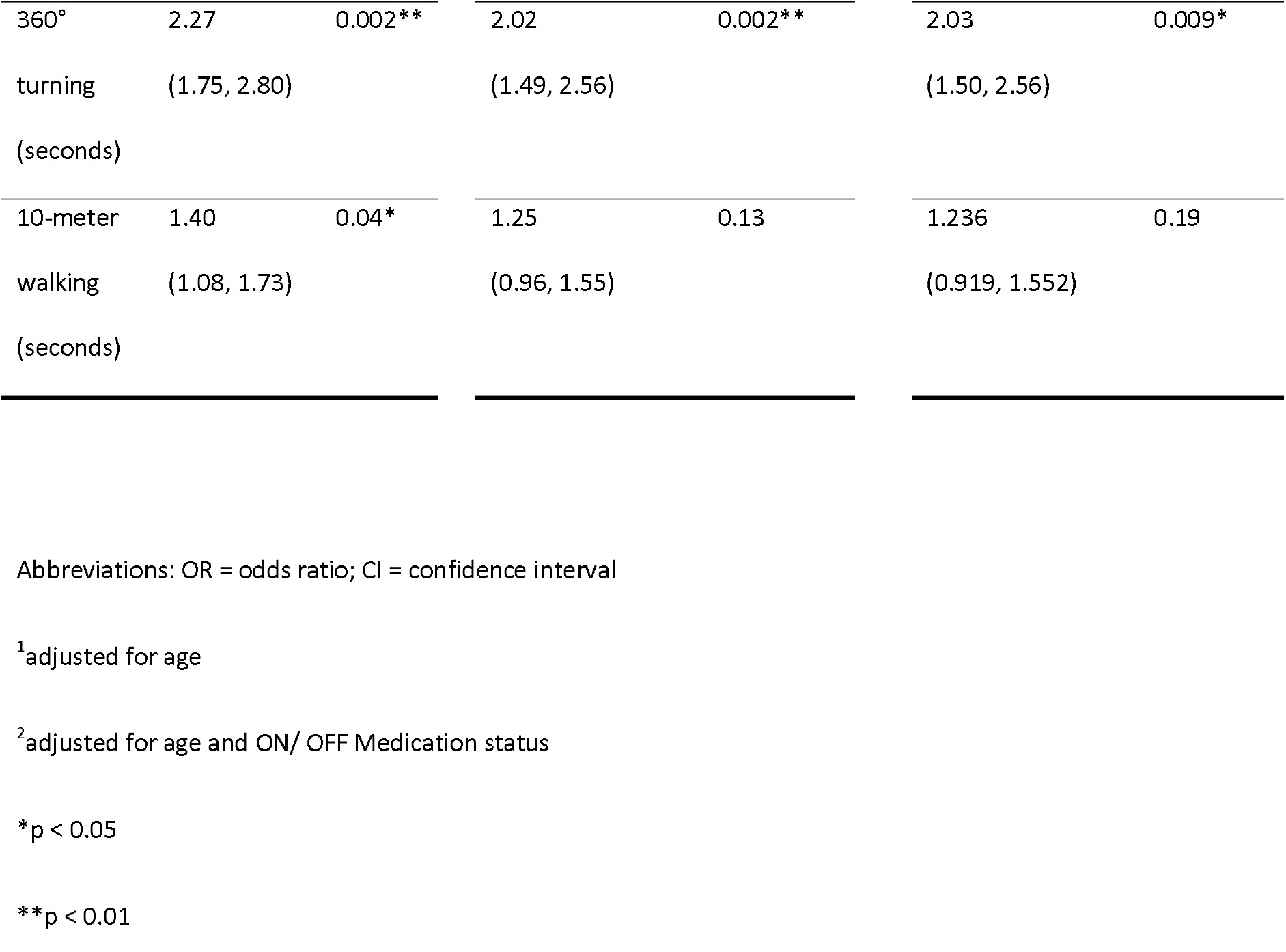
Multiple logistic regression results for clinical balance tests, adjusted for age as well as ON/OFF medication status.

### ROC Analysis

Receiver-Operating Characteristic (ROC) analysis was used to evaluate the classification accuracy for the balance measures “tandem stand duration” and “360° turning”. Since our goal was to identify patients with the PIGD subtype and imbalanced sample sizes can introduce bias into performance of the classifier, four patients of the “indeterminate” group were omitted for the next steps of the analysis.

ROC curves for tandem stand duration and 360° turning showed very high level of discrimination ability with AUCs of 86.6% (tandem stand duration) and 88% (360° turning) respectively. Classification accuracy for the combined model using tandem stand duration and 360° turning showed a high AUC of 91.4%. (Figure 2)

**Figure 2:**
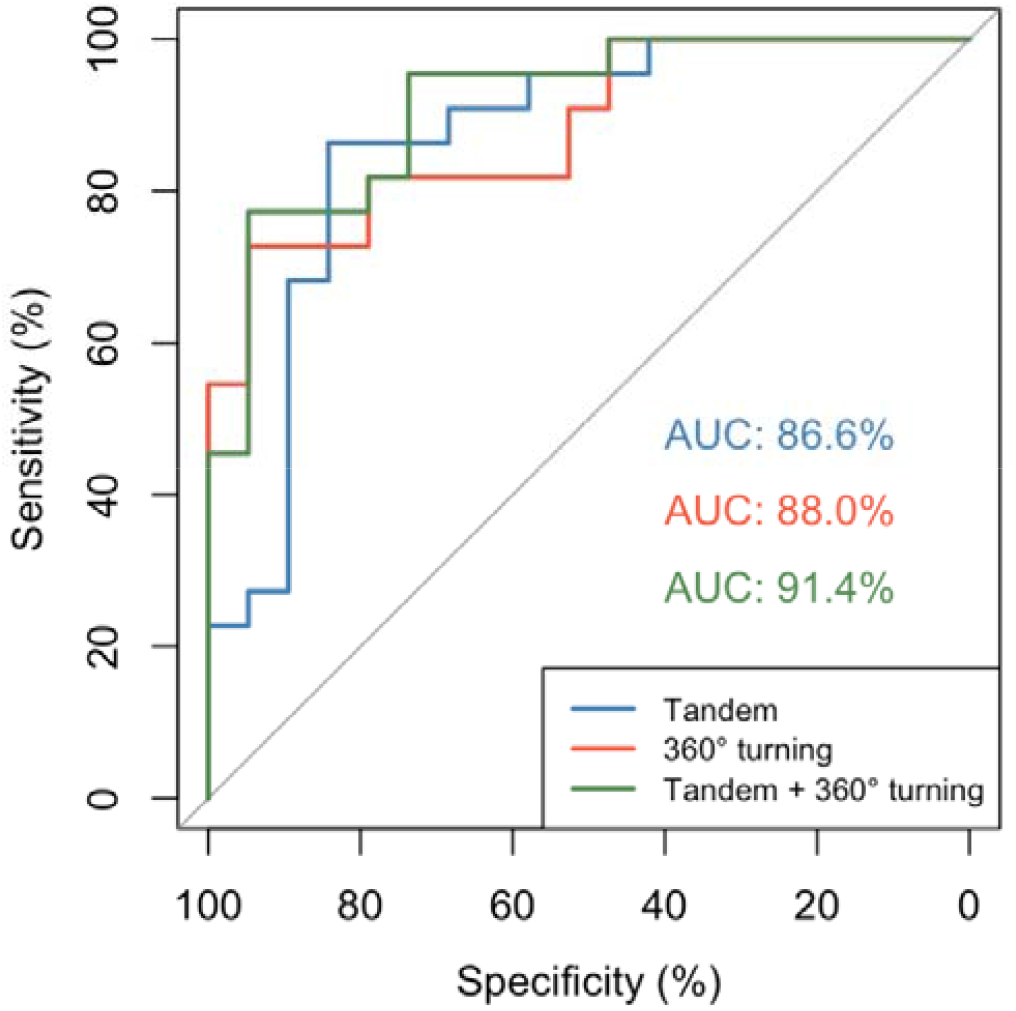
Receiver Operating Characteristic (ROC) curves for tandem stand duration, 360° turning and combined model.

Optimal threshold levels for tandem stand duration and 360° turning were selected using Youden’s index (Table 4).

**Table 4:** AUC, optimal thresholds, specificity and sensitivity (using Youden’s index) for predictive models.

|  | AUC (95% CI) | Optimal threshold | Specificity | Sensitivity |
| --- | --- | --- | --- | --- |
| Tandem stand | 86.6% (74.63 - 98.57) | 20 seconds | 84.2% | 86.4% |
| 360° turning | 88.04% (77.9 - 98.17) | 6.5 seconds | 94.7 | 72.7% |
| Tandem stand & 360° turning | 91.39% (82.91 - 99.87) | - | 94.7 | 77.3% |

## Discussion

We set out to examine tandem standing compared to other clinical balance tests as a mean to distinguish PD phenotypes. We found that a shorter duration of unassisted tandem standing as well as a longer time to turn 360° were associated with PD patients of the PIGD phenotype, while time to walk 10 meters was not a significant predictor of PD phenotype but rather depended on the age of the participants. The accuracy to predict the PD phenotype with a clinical balance test was compared using ROC curves, where both tandem standing and 360° turning showed very high accuracy with an AUC of 86.6% and 88% respectively. Combining the measures of tandem stand duration and 360° turning increased the AUC further to 91.4%. Optimal cut-off values to detect a patient with a PIGD phenotype were less than 20 seconds of tandem stand duration and more than 6.5 seconds for 360° turning.

Our data support earlier findings from Prime et al., who found time to turn 360° as well as step count of 360° turning to have a high accuracy to distinguish PIGD phenotype from TD phenotype. [13] While no difference was found in tandem walk (number of interruptions or ‘out-steps’ occurring in a ten-step tandem walk), 360° turning was able to differentiate between PIGD and TD PD subtypes with an AUC of 0.751 and 0.740 in their cohort. A possible explanation for the lower predictive accuracy might be that the patients were tested in “OFF” medication state, while our patients were tested as they presented in clinic and were mainly in “ON” state. The difference in postural stability between patients with TD and PIGD subtypes might be amplified by the effect of medication, with dopamine having a faciliatory effect on balance and gait tasks, without being the driving factor as seen in Table 3. Similar findings have been described in earlier trials, where PD patients showed no improvement in gait parameters on cholinergic medication in OFF status, while gait improvements were observed when cholinergic medication was administered alongside dopaminergic medication compared to dopaminergic medication alone. [21, 22]

Turning difficulties are observed even in mild PD patients and become especially apparent in patients with Freezing of Gait (FoG). Compared to healthy subjects, PD patients seem to turn less efficiently with exaggerated stride length reduction towards the end of a turn, increased stride width and increased percentage of double support. [23, 24] During walking, turning and not straight-line walking, is prominently under vestibular control [25], however the influence of sensory integration of vestibular, proprioceptive and visual information during turning in PD patients is still unclear. One study found PD patients were able to update their position following passive rotations in the dark (indicating preserved reflex and higher-order vestibular functioning), however the patients were all mild PD with no turning difficulty, and the use of a single test rotational velocity does not obviate a counting (time-based) strategy in completing the task. [26]

### Strengths and limitations

The main limitation to this study is the sample size of 45 patients. However, the PD phenotypes of interest (PIGD and TD) were balanced in our sample and models were adjusted accordingly to reduce the risk of overfitting.

To our knowledge this study is the first to examine tandem standing together with other balance parameters to distinguish patients with PIGD phenotypes. Tandem stand duration and time to turn 360° are accessible tests that can be done in any clinical setting, do not require special training and can identify a patient group with a high risk of falling quickly.

### Possible pathophysiological changes in PIGD subtype

Within the subtype of PIGD patients, especially patients with freezing of gait (FoG) have shown to have deficits in sensory integration with reduced ability to recruit vestibular information and depend on vestibular-visual interaction in the absence of reliable somatosensory information compared to those without FOG.[27, 28] Tandem stance reduces the base of support and leads to destabilisation in the mediolateral plane. [29] Apart from reducing mechanical stiffness that is optimized in quiet standing with hips placed above the feet, a reduction of ankle proprioceptive information due to reduced ankle range of motion in tandem stance has been discussed as a factor for increased sway. [30, 31] Based on this, we might argue that we are able to replicate the above-mentioned findings of deficient vestibular-visual signal integration in the absence of reliable proprioceptive input even through the small adaptation of proprioceptive information in tandem standing.

With the high predictive accuracy of 360° turning to identify PIGD subtypes independent of age and medication status, while no difference in 10-meter walking is observed, we suspect a distinctive pathomechanisms to be present in PIGD patients that goes beyond pure motor deficits. Failure to integrate vestibular and visual information has shown to correlate with changes of the cholinergic network with a dominance of right hemispheric cortical regions, most pronounced in Brodman area 37, the human homologue of the middle superior temporal (MST) area. [32] At the brainstem level, the mainly cholinergic pedunculopontine nucleus (PPN) has been discussed as possible structural correlate for dysfunction of sensory integration in PD patients with observed improvement of vestibular thresholds in patients with PPN deep brain stimulation. [32, 33] Decreased cholinergic activity of pedunculopontine - thalamic projections has been shown to be associated with decreased static balance in PD patients. [34] With the significant vestibular input to the pedunculopontine nucleus that has been shown to be present in primates, decreased PPN activity might explain dysfunction of sensory integration as a main factor for balance dysfunction. Further research examining PPN activity and sensory integration in PD patients, especially in the PIDG subtype is needed to understand sensory integration mechanisms in this patient group, especially of vestibular and visual information, as well as evaluation of each sensory system will be needed to better understand possible pathophysiological mechanisms and investigate future treatment options.

## Data Availability

The data supporting the findings of this study are available on request from the corresponding author. The data are not publicly available due to privacy or ethical restrictions.

## Acknowledgments

The authors would like to acknowledge the help from David Singh, who has seen and cared for many of the patients in this study in his Parkinson’s nurse clinic. We hope that he is enjoying his retirement.

## Artificial Intelligence Policy

ChatGPT, version [V 3.5], a language model developed by OpenAI (https://www.openai.com), was used for debugging and improving self-written code for statistical analysis.

